# Hyperventilation Unmasks Convergent Aperiodic EEG Vulnerability in Seizure Disorder and Schizophrenia

**DOI:** 10.64898/2026.09.19.26363459

**Authors:** Vardhan Paliwal, Alexander Moiseev, Sam M. Doesburg, Pengcheng Xi, Joel S. Winston, Mark P. Richardson, Roman Rodionov, Urs Ribary, Andrew Blaber, George Medvedev, Vasily A. Vakorin

## Abstract

Electroencephalographic (EEG) reactivity to hyperventilation probes neural responsiveness to metabolic challenge, yet no systematic study has characterized its lifespan variation. Conventional analysis further restricts inference to oscillatory dynamics, overlooking aperiodic components that reflect systems-level properties of neural population activity. We constructed brain lobe-specific lifespan reference charts for HV reactivity across oscillatory (alpha and beta power and frequency) and aperiodic (1/f slope and offset) parameters in 14,905 outpatient recordings (ages 1–88), then quantified normative deviations in seizure disorder (N=115) and schizophrenia (N=154), two conditions with mechanistically opposing excitatory-inhibitory dysfunction. Adolescence (~age 12) marked a critical inflection point: oscillatory power reactivity converged toward zero while aperiodic parameters reached peak dynamic range. As expected from opposing pathophysiology, seizure disorder and schizophrenia showed divergent oscillatory signatures, though effect sizes were modest (Cohen’s d *<* 0.2). Both disorders, however, converged on hyporeactive aperiodic offset with large effect sizes (Cohen’s d *>* 0.5 across all lobes), indicating shared impairment in modulating broadband population activity under metabolic stress. These findings establish the first lifespan reference charts for HV reactivity and reveal a dissociation between oscillatory and aperiodic parameters: aperiodic offset captures shared metabolic vulnerability that transcends diagnostic boundaries and emerges as a candidate transdiagnostic biomarker.

## Introduction

Probing electroencephalographic (EEG) reactivity to hyperventilation (HV) represents a standard clinical procedure during routine EEG to assess brain function by observing changes in cerebral electrical activity during voluntary, rapid, deep breathing. Although HV has remained a standard neurophysiological assessment for decades, recent research reveals that it functions not merely as a seizure provocation technique, but as a controlled physiological challenge that probes how neural networks respond to metabolic perturbation (Patel and Maulsby (1987); Rana et al. (2023)). HV reduces arterial carbon dioxide, inducing cerebral vasoconstriction and respiratory alkalosis that collectively shift neuronal networks toward increased excitability by disrupting excitatory-inhibitory (E/I) balance, defined as the dynamic equilibrium between excitatory and inhibitory neuronal activity governing neural oscillations, network stability, and information processing efficiency (Yizhar et al. (2011)). This complexity positions HV as a metabolic stress test capable of exposing vulnerabilities in neural system regulation inaccessible to resting state measures.

The brain’s EEG response to this metabolic challenge can be decomposed into two distinct levels of neural organization. Oscillatory parameters, including alpha and beta band rhythms, capture the activity of specific neural circuits operating at characteristic timescales: alpha reflects thalamocortical inhibitory dynamics during wakeful rest, while beta rhythms represent localized cortico-cortical excitation-inhibition interactions (Buzsaki and Draguhn (2004)). Aperiodic 1/f spectral parameters capture systems-level properties: the 1/f spectral slope serves as a proxy of overall E/I balance (Gao et al. (2017); Kluger et al. (2023)), while the aperiodic offset reflects total broadband power correlating directly with neural population spiking rates (Manning et al. (2009); Miller et al. (2014); Voytek and Knight (2015)). These components can be independently affected by pathology, with some disorders showing purely oscillatory alterations while others show exclusively aperiodic changes (Pani et al. (2022)). This dissociation has direct relevance for HV: prior studies have characterized only oscillatory responses to HV, reporting established alpha modulation patterns (Yang et al. (2025); Gibbs et al. (1943)) but leaving beta modulation entirely uncharacterized, and no study has examined aperiodic reactivity to HV.

Age is a critical determinant of HV response. In a landmark comparative study, Nadarajah and colleagues found that HV makes absence seizure capture 2.4 times more likely in children compared to those who did not undergo HV, with no significant benefit in adults (Nadarajah et al. (2024)). This stark age difference, noted in historical reviews (Patel and Maulsby (1987); Khachidze et al. (2021)), likely relates to developmental changes in cerebrovascular responsiveness and inhibitory system maturation. Studies comparing EEG reactivity to HV across age groups found declining alpha power and frequency reactivity with increasing age (Konishi (1987); Ponomareva et al. (2012)). Yet no systematic study has mapped continuous EEG spectral parameter reactivity to HV across the human lifespan. Without normative reference data, clinicians cannot distinguish pathological HV responses from age-typical variation.

Patient pathology constitutes another critical factor, with different neuropsychiatric disorders exhibiting distinct patterns of E/I balance dysfunction that should produce divergent responses to physiological provocation. Seizure disorders show E/I balance shifts toward excessive excitation (Fritschy (2008)), and HV represents a well-established method for activating epileptiform discharges, particularly in generalized epilepsies (Rana et al. (2023); Alghamdi et al. (2021)). Comparing patients with epilepsy to controls reveals higher alpha power reactivity to HV, consistent with underlying hyperexcitable states (Gibbs et al. (1943)). Conversely, schizophrenia spectrum disorders demonstrate altered E/I balance through loss of inhibitory neuronal activity (Lewis et al. (2005)). Bose and colleagues found significantly less EEG power change during HV in schizophrenia (Bose et al. (2016)), and only 3% of patients with schizophrenia showed EEG slowing during HV compared to 55% of controls, indicating increased central nervous system resistance to HV consistent with the general physiological unresponsiveness of this disorder (RUBIN (1942)). Despite these opposing mechanisms, epidemiological evidence reveals shared susceptibility between the two disorders: bidirectional associations show approximately 6-fold higher epilepsy risk in schizophrenia and 8-fold higher schizophrenia risk in epilepsy (Chang et al. (2011); Cascella et al. (2009)), and recent genomic studies have identified over 40 shared genetic risk loci (Karadag et al. (2023)). This convergence raises the possibility that despite opposing circuit-level manifestations, these disorders may share deeper systems-level vulnerabilities that could be revealed by appropriate physiological probes.

This study addresses two aims: (1) to model EEG spectral parameter reactivity to HV across the human lifespan, encompassing oscillatory and aperiodic parameters; and (2) to quantify how seizure disorder and schizophrenia alter these reactivities relative to normative trajectories. We hypothesized that HV reactivity would show systematic age-related changes reflecting cortical network maturation, with greater responses during childhood when neural plasticity peaks. Given their opposing E/I dysfunction, we further hypothesized that seizure disorder and schizophrenia would manifest opposing oscillatory reactivity patterns. We also characterized aperiodic parameters, for which prior literature offered no clear directional predictions.

## Methods

### Patient Demographics

We analyzed routine clinical EEG recordings acquired from four public hospitals within the Fraser Health Authority, British Columbia, Canada, between January 2010 and December 2018. No new human subjects were recruited or tested. The original data were collected under ethical approval by Simon Fraser University and Fraser Health Authority on 1^*st*^ April 2022 (protocol number: H18-02728). All data were anonymized/de-identified prior to analysis. We categorized recordings according to patient status (outpatient or inpatient) following the electronic health records. Not all routine clinical EEG tests included HV. We selected only EEGs with HV, without any bias. Thus, our study cohort reflected a culturally and socioeconomically diverse patient population.

Our control group comprised outpatient EEGs from individuals visiting hospitals for EEG evaluation but not requiring admission (N = 14,905), with balanced sex representation. Outpatient EEGs were selected for reference model construction because they provide comprehensive lifespan coverage from infancy through advanced age — coverage that dedicated healthy control cohorts cannot practically achieve. While this population does not constitute a purely healthy control group, the large sample size (N = 14,905) enabled averaging of individual pathological effects when modeling population-level age trajectories, yielding clinically relevant reference standards representative of general clinical populations against which clinical groups can be systematically compared. This approach prioritizes translational applicability to routine clinical practice over idealized but practically unachievable lifespan coverage from strictly healthy cohorts.

Our clinical groups comprised inpatients who spent at least one night receiving acute hospital care, excluding emergency department-only visits. We categorized inpatient EEG into Case Mix Groups (CMGs) defined by CIHI based on International Statistical Classification of Diseases and Related Health Problems, 10th Revision, Canada (ICD-10-CA). This categorization yielded 21 CMG groups, each representing a Major Clinical Category (MCC). For this study, we selected MCC-17 (Mental Diseases and Disorders) as our primary inpatient category. Within this category, we identified the two most prevalent diagnostic groups: seizure (N = 115) and schizophrenia disorders (N = 154). The schizophrenia cohort encompassed conditions ranging from mild schizotypal disorder to severe schizophrenia manifestations, representing the full clinical spectrum. These groups were selected because they represent mechanistically distinct and opposing forms of E/I balance dysfunction: seizure disorder characterized by network hyperexcitability and schizophrenia by network rigidity and EEG Reactivity to Hyperventilation, YEAR, Volume XX, Issue x 3 inhibitory deficits, making them ideally suited to test our hypothesis that opposing pathophysiological states would produce divergent HV reactivity signatures. Furthermore, these disorders have the most established, albeit limited, prior literature on EEG responses to HV, providing a mechanistic framework against which our spectral findings could be interpreted. Remaining diagnostic groups within the Mental Disorders MCC were insufficiently powered for reliable trajectory modeling and were therefore excluded. Demographics for all three EEG cohort groups are summarized in Table 1, with age distributions visualized in Fig. 1.

**Table 1.**
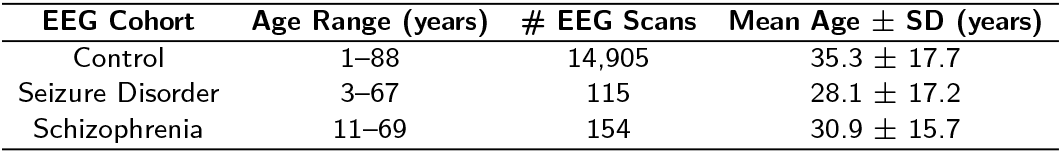
Demographics of EEG Cohorts.

**Figure 1.**
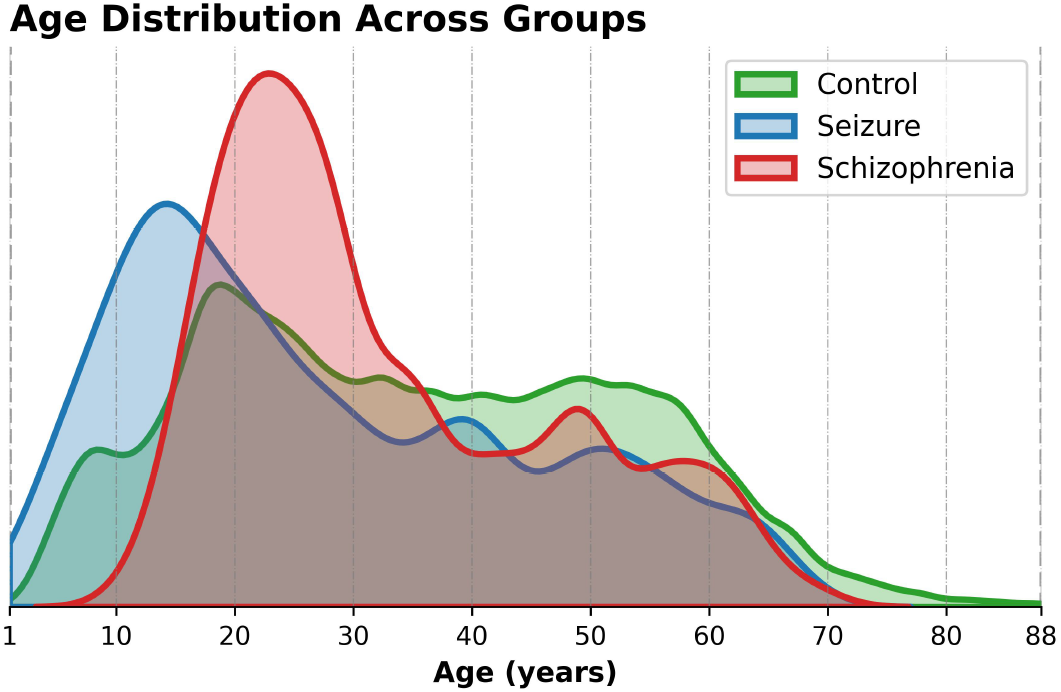
Age distribution of EEG cohorts present in the analysis. Kernel density plots displaying the age distribution for control group (green curve; *N* = 14,905; age: [1, 88] years), seizure group (blue curve; *N* = 115; age: [3, 67] years), and schizophrenia group (red curve; *N* = 154; age: [11, 69] years).

### EEG Processing

EEG was recorded using standardized hardware and protocols across all four clinical sites. Each recording station utilized a Natus Xltek EEG32U amplifier coupled with gold-cup electrodes positioned according to the International 10-20 system. We collected data from 20 standardized electrode locations: FP1, FPZ, FP2, F3, F4, F7, F8, FZ, T3, T4, T5, T6, C3, C4, CZ, P3, P4, PZ, O1, and O2. Original sampling frequencies were maintained at either 500 Hz or 512 Hz depending upon the acquisition site.

We converted raw EEG from native Natus proprietary format to European Data Format (EDF) and anonymized them using the PyEDFlib library (Nahrstaedt et al. (2020)) in Python. Signal preprocessing employed zero-phase, overlap-add finite impulse response bandpass filtering (0.5-55 Hz) with Hamming window implementation via the MNE-Python library (Gramfort et al. (2013)). This frequency range preserved canonical neurophysiological rhythms while attenuating artifacts above the 60 Hz powerline frequency. We resampled all recordings to 256 Hz for temporal standardization across the dataset. Artifact removal protocols eliminated flat signal intervals (defined as digital zeros with peak-to-peak amplitude *<*1e-6) and photic stimulation epochs. The final preprocessed dataset yielded two data segments per participant: 6-minute clean RS EEG recordings and variable-length HV episodes, both consisting of the same 20-channels.

### EEG Spectral Parameter Extraction

We reconstructed EEG source activities for both clean RS segments (Moiseev et al. (2025)) and HV episodes using 148 cortical regions of interest (ROIs), with 74 ROIs per hemisphere, according to the Destrieux cortical atlas (Destrieux et al. (2010)). Source reconstruction employed a standardized fsaverage template MRI from FreeSurfer with standard electrode positions for the 10-20 montage, consistent with established practice for large-scale retrospective clinical EEG cohorts where individual MRIs are unavailable. This template was applied uniformly across all age groups. While standard for large-scale retrospective cohorts, uniform template application may introduce greater spatial uncertainty in younger participants whose head geometry deviates more substantially from the adult average, a consideration addressed in the Limitations. Each ROI was categorized into one of six cortical lobes: insular, limbic, frontal, occipital, parietal, and temporal lobes, following anatomical definitions provided in the Destrieux parcellation. Forward solutions (lead field matrices) were computed for each cortical source using the MNE-Python library. Forward modeling employed a three-layer boundary element method (BEM) based on individual patient head models, with tissue conductivity values: inner skull (*σ* = 0.3 S/m), outer skull (*σ* = 0.006 S/m), and scalp (*σ* = 0.3 S/m) (Hillebrand and Barnes (2005); Frost (1972); Sekihara and Nagarajan (2008)). Neural source activity estimation utilized a scalar linearly constrained minimum variance beamformer (Robinson (1999); Sekihara and Nagarajan (2008)). Beamformer weights were computed to minimize output variance while maintaining unit gain for the source of interest, effectively suppressing contributions from other brain regions and external noise sources. To correct for depth-dependent biases inherent in beamformer solutions, we normalized reconstructed time series by dividing by the root mean square (RMS) amplitude of the projected sensor noise covariance. This normalization yields dimensionless pseudo-Z scores, where values represent signal-to-noise ratio at each source location, enabling fair comparison across cortical regions at different depths.

We computed power spectral density for each source time series using Welch’s method (Welch (1967)). Spectral analysis covered 1-55 Hz with 0.5 Hz resolution, yielding 109 frequency bins. The final dataset comprised spectral power estimates for 148 cortical sources across 109 frequency points for both RS and HV segments.

From source-reconstructed EEG spectral power, we extracted spectral parameters using the spectral parameterization toolbox (Donoghue et al. (2020)). This method decomposed EEG spectral power into aperiodic (1/f) components and Gaussian distributions modeling spectral peaks associated with oscillatory brain rhythms. The 1/f aperiodic component was characterized by two parameters: slope, quantifying the rate of spectral power decay with increasing frequency, and offset, quantifying the vertical shift of the power spectrum reflecting broadband spectral power magnitude. Gaussian peaks represented distinct oscillatory rhythms in the power spectrum.

We defined alpha rhythms within 7-13 Hz; when multiple peaks appeared within this range, we selected the highest power peak as alpha frequency with corresponding power as alpha power (arbitrary units). Similarly, we defined beta rhythms within 13-30 Hz; when multiple peaks appeared, we selected the highest power peak as beta frequency with corresponding power. This method isolated alpha and beta oscillations from 1/f spectrum baseline activity. We computed alpha/beta peak frequencies and powers and aperiodic slope and offset individually for each ROI, then performed lobe-wise averaging (median) across ROIs within a lobe to obtain representative values for each lobe.

### Spectral Reactivity Quantification, Normative Modeling, and Clinical Group Comparison

EEG spectral parameter reactivity to HV was defined as the difference in spectral parameter estimates between the HV and RS conditions. For each EEG spectral parameter, reactivity was calculated as: Reactivity = (*HV − RS*)_*Parameter*_. This formulation quantifies the neural response to HV relative to baseline brain activity.

Using data from control group, we generated scatterplots showing EEG reactivity across the lifespan, stratified by six anatomically defined brain lobes (insular, limbic, frontal, occipital, parietal, and temporal). To extract developmental trajectories from the data, we applied locally-weighted scatterplot smoothing (LOWESS), a non-parametric regression method implemented via the Statsmodels library (Seabold and Perktold (2010)). LOWESS estimates non-linear age trends while removing the influence of outliers, providing smoothed reference trajectories.

To evaluate alterations in EEG reactivity to HV in two clinical populations, we compared the EEG reactivity trajectories of individuals with seizure disorder and schizophrenia to those control group. For each clinical group, we focused on the age range defined by the 5th and 95th percentile of that group’s age distribution: 6.0-64.0 years for seizure disorder and 16.6-60.3 years for schizophrenia. Control group trajectories were modeled across the full combined 5th and 95th percentile age span (6.0-60.4 years). All clinical and control group trajectories were smoothed using the same LOWESS procedure.

We categorized clinical deviations in reactivity patterns as hyperreactive or hyporeactive relative to the control reference. For control trajectories with positive values (*>*0), clinical values exceeding the control curve were classified as hyperreactive, whereas values below the curve were deemed hyporeactive. Conversely, when control trajectories were negative (*<*0), clinical values above the curve were interpreted as hyporeactive, and values below as hyperreactive.

To quantify individual-level deviations, each clinical EEG’s reactivity values were projected onto the corresponding age-matched control trajectory to derive deviation scores for each brain lobe. These scores were then aggregated to characterize group-level effects. We assessed statistical significance using one-sample t-tests comparing mean deviations to zero. Additionally, we computed effect sizes using Cohen’s d to evaluate the magnitude of deviation. To account for multiple comparisons, p-values were adjusted using the Benjamini-Hochberg procedure, applying a false discovery rate (FDR) threshold of 10%. Statistical significance was defined as *p <* 0.1.

Effect size (Cohen’s *d*) was the primary metric of clinical deviation magnitude, as it quantifies the practical significance of group differences independently of sample size. Consistent with ASA guidelines on statistical inference (Wasserstein and Lazar (2016); Wasserstein et al. (2019)), p-values were treated as continuous descriptive statistics and interpreted in conjunction with effect sizes rather than as binary arbiters of significance: a p-value threshold does not demarcate findings that are real from those that are not, and should not be used as such. Following Benjamini-Hochberg false discovery rate correction across brain lobes and spectral parameters, *p <* 0.1 was used as a secondary screening heuristic to organize reported findings; it governed what was reported, not what was concluded. The magnitude and direction of Cohen’s d remains the basis for all clinical interpretations.

## Results

### Alpha Power Reactivity to HV Shows Developmental Convergence but Clinical Divergence

We first characterized alpha power reactivity, as alpha power represents the most extensively studied EEG parameter in the context of HV and provides the most direct test of our normative modeling approach. Alpha power reactivity was negative in childhood, reflecting suppression of alpha power during HV, and increased steadily toward zero by adolescence onset (~12 years), plateauing thereafter across all brain lobes (Fig. 2**A**,**B**). This developmental convergence toward zero reactivity indicates that the alpha power response to metabolic challenge diminishes as the brain matures.

**Figure 2.**
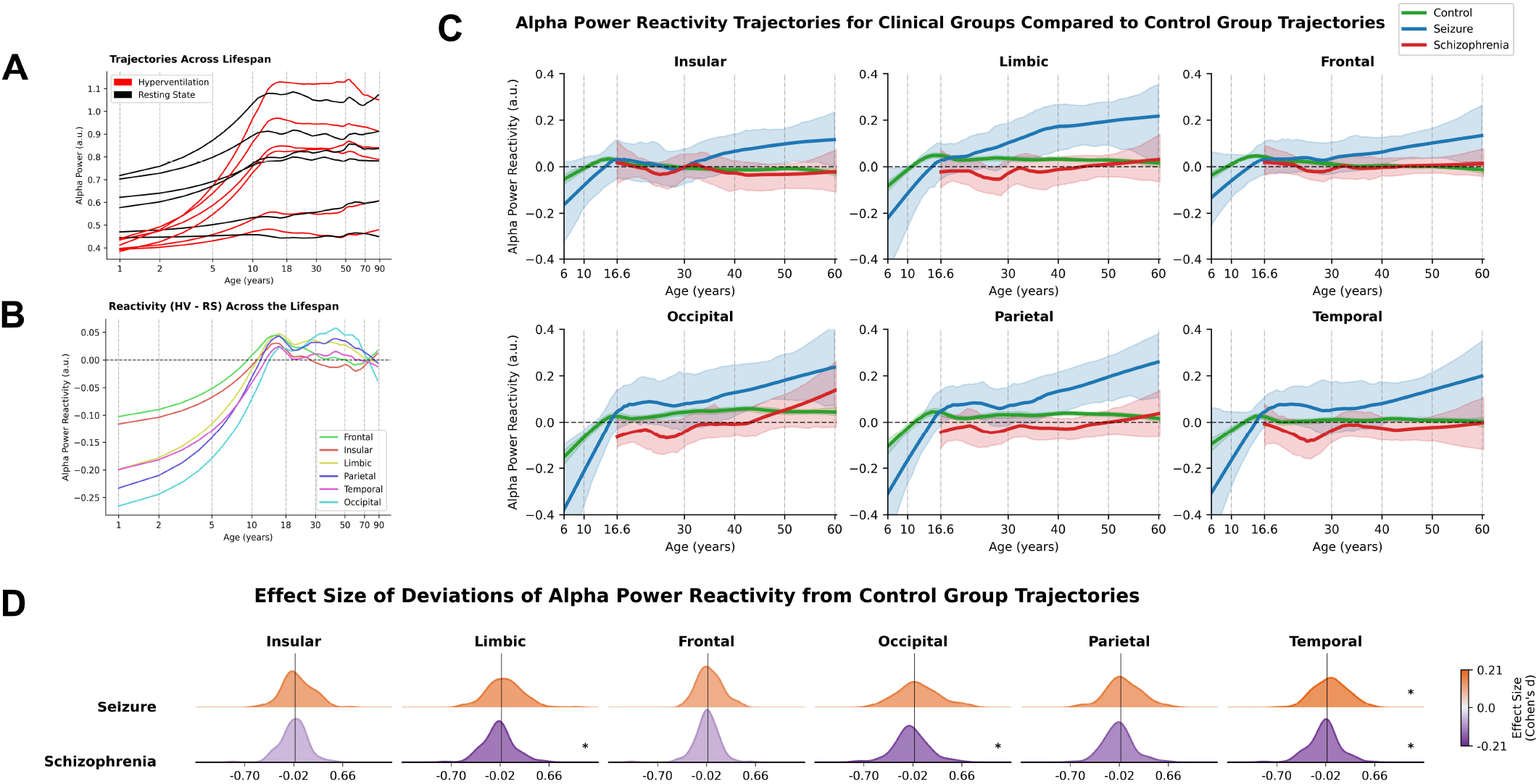
Effects of age and brain pathology on alpha power reactivity: Age-related normalization and divergent pathological effects. Panels A-D characterize alpha power reactivity across lifespan and clinical groups, stratified by brain lobes. **(A)** Visual comparison of alpha power changes throughout lifespan for HV and RS EEG conditions. Age is represented on natural logarithmic scale to emphasize developmental changes. Alpha power in children is lower during HV compared to RS across all six brain lobes. With increasing age, alpha powers converge across the two conditions during adolescence, and remains stable thereafter. **(B)** Lifespan reference charts for alpha power reactivity to HV. Age is shown on natural logarithmic scale. Alpha power reactivity begins with negative values in children, increasing until adolescence onset (age ~ 12 years), then plateaus around zero for remaining lifespan. The pattern is consistent across brain lobes. **(C)** Visual comparison of alpha power reactivity trajectories for 5th-95th percentile age range for control, seizure, and schizophrenia groups. Trajectories are compared on a linear age scale. Seizure group trajectories are higher than control group; schizophrenia trajectories are lower. Since control trajectory values are either positive or zero in this age range, this means that seizure group shows hyperreactivity while schizophrenia shows hyporeactivity. **(D)** Effect size (Cohen’s d) of distribution of deviations in individual alpha power reactivities in clinical groups. Orange distributions show positive effects (values above reference); violet distributions show negative effects (values below reference). While the effects are consistent across brain lobes, both seizure disorder and schizophrenia groups show weak effect sizes (Cohen’s *d <* 0.2).

Against this normative reference, the two clinical groups diverged in the predicted directions. Seizure disorder showed hyperreactivity with excessive alpha enhancement during HV, while schizophrenia showed hyporeactivity with blunted alpha enhancement relative to controls, across all brain lobes (Fig. 2**C**). Effect sizes were modest (Cohen’s *d <* 0.2) but directionally consistent with opposing pathophysiology, with positive effects for seizure disorder and negative effects for schizophrenia (Fig. 2**D**).

### Age-Related Enhancement of Alpha Frequency Reactivity is Maintained in Seizure Disorder but Diminished in Schizophrenia

We next examined alpha frequency reactivity, which reflects thalamocortical timing rather than amplitude and constitutes a dissociable feature of the alpha rhythm. Alpha frequency reactivity increased continuously across the lifespan: negative in childhood reflecting alpha slowing during HV, crossing zero in young adulthood, and becoming positive thereafter as HV began to accelerate rather than slow alpha frequency (Fig. 3**A**,**B**). This monotonic trajectory suggests that the thalamocortical timing response to metabolic challenge undergoes a prolonged developmental recalibration, distinct from the adolescence-limited convergence seen in alpha power.

**Figure 3.**
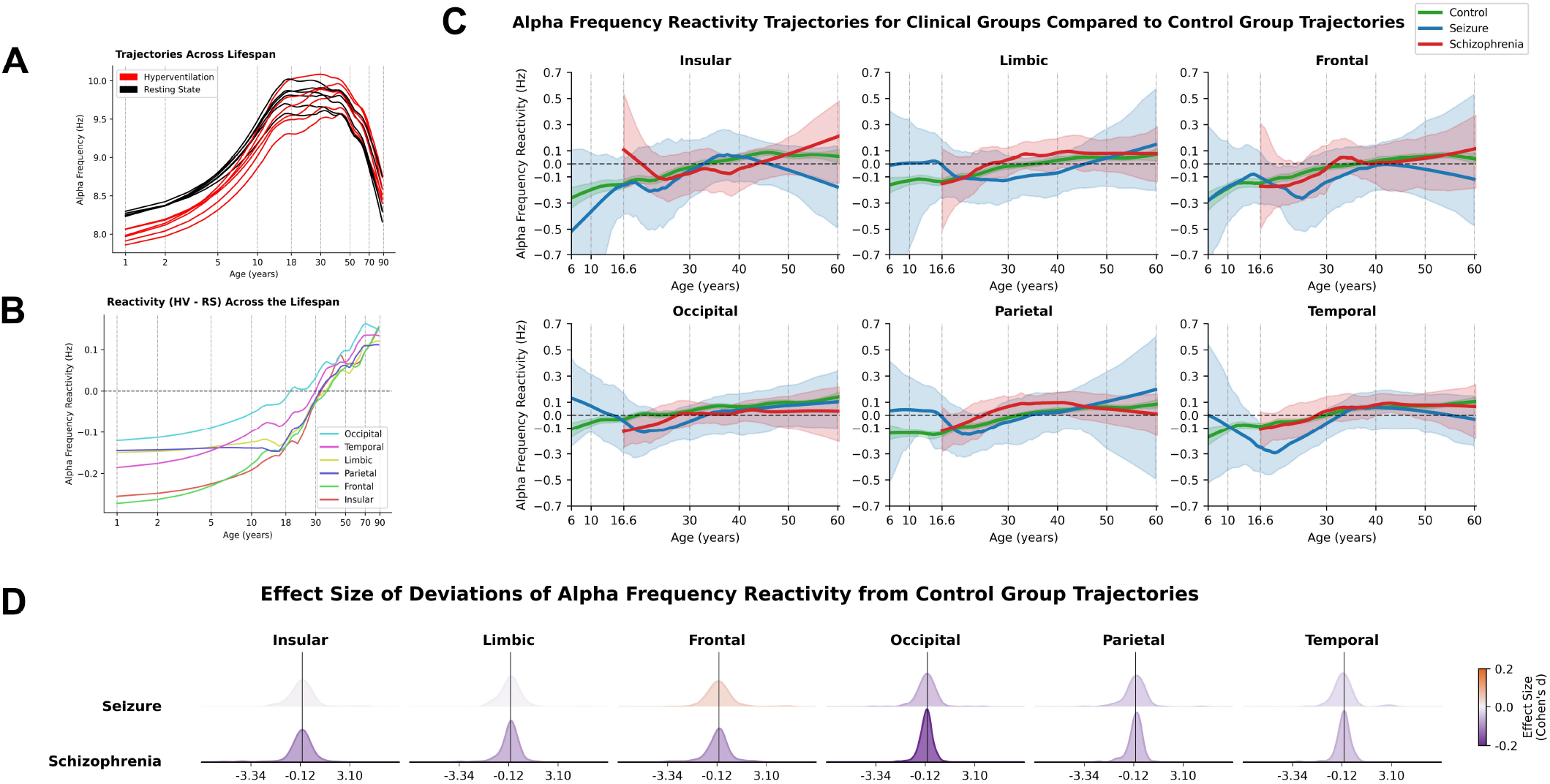
Effects of age and brain pathology on alpha frequency reactivity: Age-related increase in reactivity values that exhibit preserved response in seizure disorder but blunted response in schizophrenia. Panels A-D characterize alpha frequency reactivity across lifespan and clinical groups, stratified by brain lobes. **(A)** Visual comparison of alpha frequency changes throughout lifespan for HV and RS conditions. Age is shown on natural logarithmic scale. Alpha frequency in children is lower during HV compared to RS. Values converge around young adulthood, followed by alpha frequency becoming higher during HV in later years. **(B)** Lifespan reference charts for alpha frequency reactivity to HV. Age is shown on logarithmic scale. Alpha frequency reactivity increases continuously throughout lifespan, with negative values in childhood crossing zero around young adulthood, and becoming positive in later years. The pattern is consistent across brain lobes. **(C)** Visual comparison of alpha frequency reactivity trajectories for control, seizure, and schizophrenia groups within 5th-95th percentile age ranges on linear age scale. Clinical group trajectories appear similar to controls on visual inspection. **(D)** Effect size (Cohen’s d) of distribution of deviations of individual alpha frequency reactivity. Violet distributions show negative effects (values below reference). Seizure group shows near-zero effect sizes indicating preserved alpha frequency reactivity while schizophrenia shows weak negative effect sizes implying hyporeactivity with blunted alpha frequency enhancement.

Comparing alpha frequency reactivity for clinical groups with normative references revealed that both clinical groups showed reactivity values close to control group (Fig. 3**C**). Seizure disorder showed near-zero effect sizes (Cohen’s *d <* 0.1), indicating preserved alpha frequency modulation during HV. Schizophrenia showed modest negative effect sizes indicating hyporeactivity with blunted alpha frequency acceleration relative to controls (Fig. 3**D**).

### Beta Power Reactivity to HV Shows Developmental Convergence but Clinical Divergence

We extended the analysis to beta power reactivity to determine whether the developmental and clinical patterns observed for alpha power generalize across oscillatory parameters. Beta power reactivity followed a bi-phasic developmental trajectory: negative in early childhood, rising through a positive inflection point at adolescence onset (~12 years), then declining back toward zero by late adolescence and remaining stable thereafter across all brain lobes (Fig. 4**A**,**B**). The transient adolescent positive reactivity, absent in alpha power, indicates that beta and alpha power reactivity follow distinct developmental trajectories. The clinical reactivity pattern mirrored alpha power: seizure disorder showed hyperreactivity and schizophrenia showed hyporeactivity, with modest but directionally consistent effect sizes (Cohen’s *d <* 0.2) across all lobes (Fig. 4**C**,**D**).

**Figure 4.**
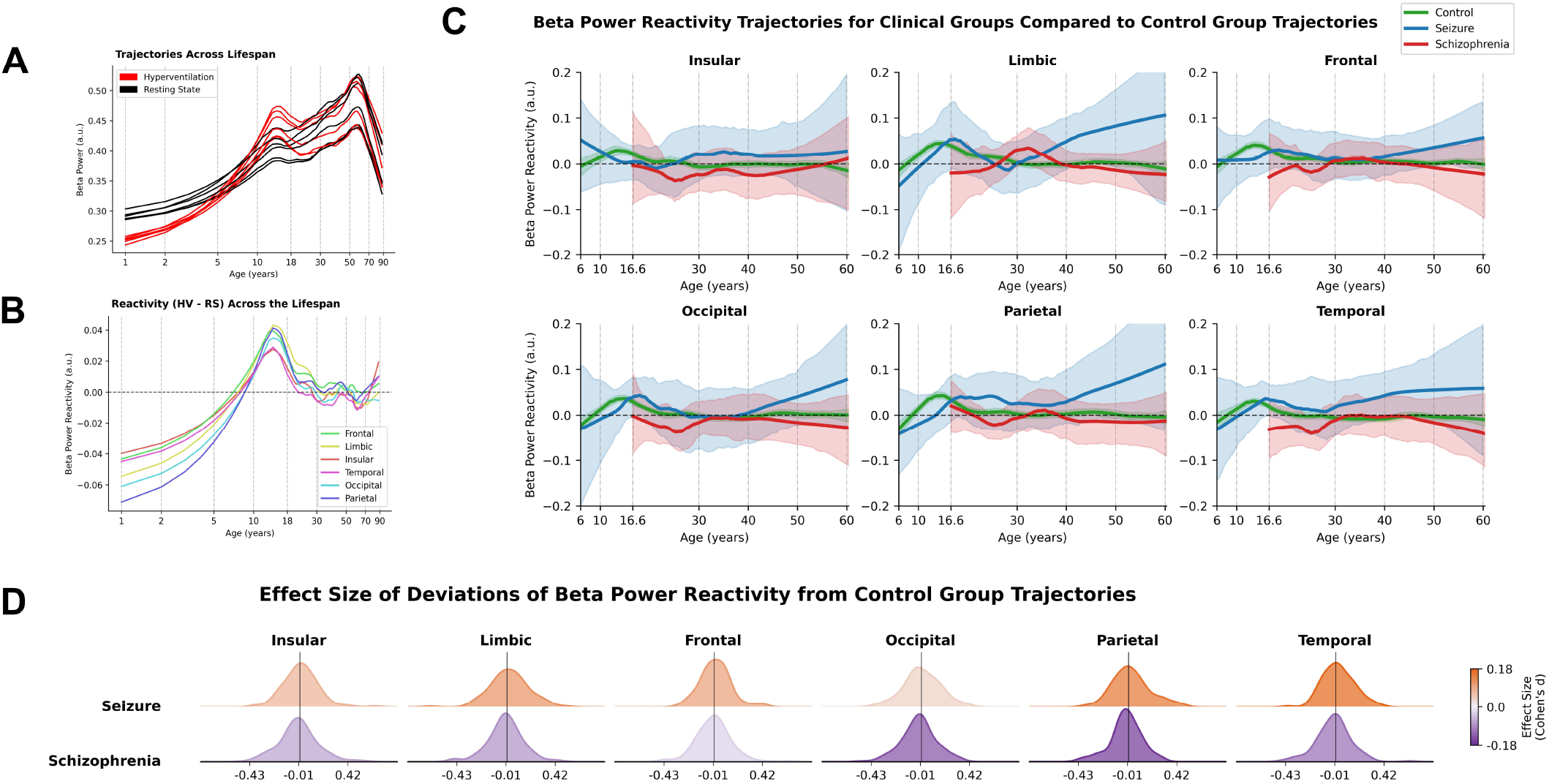
Effects of age and brain pathology on beta power reactivity: Age-related normalization and divergent pathological effects. Panels A-D characterize beta power reactivity across lifespan and clinical groups, stratified by brain lobes. **(A)** Visual comparison of beta power changes throughout lifespan for HV and RS conditions. Age is represented on natural logarithmic scale. Beta power is lower during HV compared to RS in children. During late childhood to adolescent years, beta power becomes higher during HV, followed by a convergence during adulthood. **(B)** Lifespan reference charts for beta power reactivity. Age is shown on logarithmic scale. Beta power reactivity is negative in children. It increases until adolescence onset (~12 years), becomes positive reaching an inflection point, before decreasing back to zero by late adolescence where it plateaus. The effect is consistent across brain lobes. **(C)** Visual comparison of beta power reactivity trajectories for control, seizure, and schizophrenia cohorts within 5th-95th percentile age ranges on linear age scale. Seizure group demonstrate higher reactivity values while schizophrenia shows lower reactivity values compared to control group. Since control values are either zero or positive, this means that seizure group shows hyperreactivity (excessive beta power enhancement) while schizophrenia shows hyporeactivity (blunted beta enhancement). **(D)** Effect sizes of individual deviation distributions of beta power reactivity. Orange distributions indicate positive effects; violet distributions indicate negative effects. Both groups show weak effect sizes (Cohen’s *d <* 0.2): seizure disorder show positive effect size confirming hyperreactivity while schizophrenia shows negative effect size confirming hyporeactivity. The effect is consistent across brain lobes.

### U-Shaped Developmental Beta Frequency Reactivity Trajectory is Maintained in Seizure Disorder but Blunted in Schizophrenia

Paralleling our approach for alpha, we next examined beta frequency reactivity separately from beta power to provide a complete characterization of beta rhythm modulation during HV. Beta frequency reactivity followed a U-shaped trajectory across the lifespan: consistently negative reflecting beta slowing during HV, reaching its most negative values in adolescence before partially recovering through adulthood (Fig. 5**A**,**B**). In terms of clinical group reactivity, seizure disorder showed near-zero effect sizes (Cohen’s *d <* 0.1), indicating preserved beta frequency modulation during HV. Schizophrenia showed weak positive effect sizes indicating hyporeactivity with blunted beta frequency slowing relative to controls, consistent with the selective frequency modulation impairment observed for alpha frequency in the same group (Fig. 5**C**,**D**).

**Figure 5.**
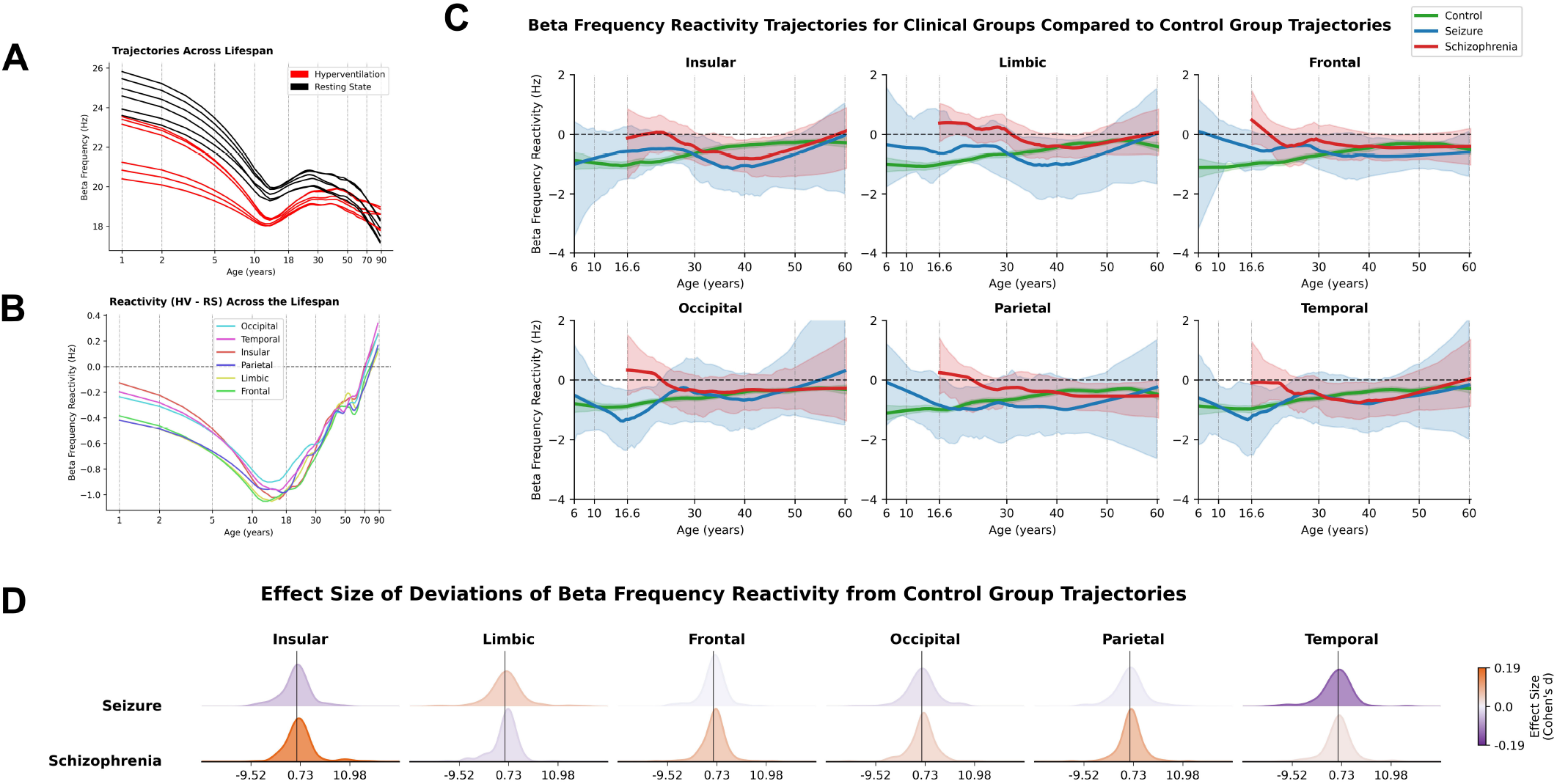
Effects of age and brain pathology on beta frequency reactivity: Age-related U-shaped trajectory is maintained in seizure disorder but diminished in schizophrenia. Panels A-D characterize beta frequency reactivity across lifespan and clinical groups, stratified by brain lobes. **(A)** Visual comparison of beta frequency throughout lifespan for HV and RS conditions. Age is shown on logarithmic scale. Beta frequency remains consistently lower during HV compared to RS throughout the lifespan. **(B)** Lifespan reference charts for beta frequency reactivity to HV. Age is shown on logarithmic scale. Beta frequency reactivity starts negative in children, decreases further reaching its most negative values during adolescence, then increases in adulthood while remaining negative. This characteristic U-shaped trajectory was consistent across brain lobes. **(C)** Comparison of beta frequency reactivity trajectories for control, seizure, and schizophrenia groups. Trajectories are compared for 5th-95th percentile age range of clinical groups and are shown on linear age scale. Clinical trajectories appear on similar levels compared to controls on visual inspection. **(D)** Effect size (Cohen’s d) of distribution of deviations in individual beta frequency reactivities in clinical groups. Orange distributions show positive effects (values above reference). Effect sizes reveal preserved modulation in seizure disorder (near-zero effect size) but hyporeactivity in schizophrenia (weak but positive effect sizes indicating blunted frequency reduction).

Across all four oscillatory parameters, a consistent clinical architecture emerged: seizure disorder showed hyperreactive power responses while schizophrenia showed hyporeactive responses, and frequency modulation was preserved in seizure disorder but blunted in schizophrenia. Effect sizes were uniformly weak (Cohen’s *d <* 0.2) across all parameters and brain lobes. These directionally consistent but modest deviations confirm that the normative framework captures disorder-relevant variation aligned with known pathophysiology, while also establishing that oscillatory parameters alone cannot robustly identify shared vulnerability across these disorders. We therefore turned to aperiodic parameters, which reflect systems-level properties of neural population dynamics rather than the activity of specific oscillatory circuits.

### Inverted U-Shaped Aperiodic Slope Reactivity Between Negative Childhood and Adolescent Phases Shows Opposing Clinical Modulation

Having fully characterized reactivity of periodic oscillatory parameters, we next next characterized the aperiodic 1/f component reactivity, beginning with aperiodic slope reactivity, which reflects population-level E/I balance rather than circuit-specific oscillatory dynamics. Aperiodic slope reactivity followed an inverted U-shaped trajectory: negative in childhood, rising to a positive peak around adolescence onset (~12 years), then declining through adulthood to stable negative values (Fig. 6**A**,**B**). The adolescent peak coincides with the inflection points observed across oscillatory parameters, consistent with a shared developmental turning point in HV reactivity across spectral dimensions.

**Figure 6.**
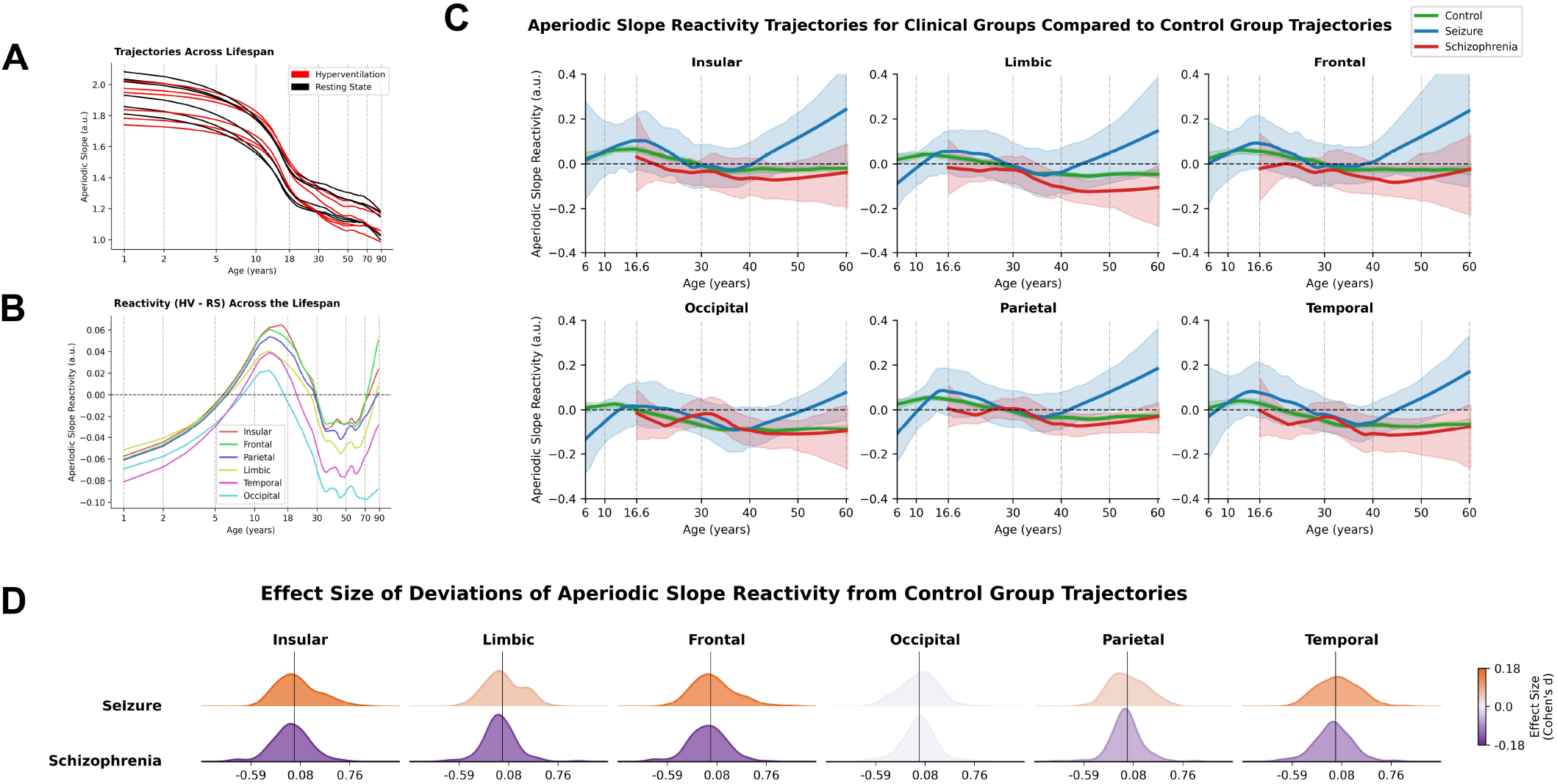
Effects of age and brain pathology on aperiodic slope reactivity: Inverted-U shaped developmental trajectory shows divergent patterns in brain pathology. Panels A-D characterize aperiodic slope reactivity across lifespan and clinical groups, stratified by brain lobes. **(A)** Visual comparison of aperiodic slope throughout lifespan for HV and RS conditions. Age is shown on logarithmic scale. Aperiodic slope in children is lower (flatter) during HV compared to RS. **(B)** Lifespan reference charts for aperiodic slope reactivity to HV. Aperiodic slope reactivity demonstrates an inverted-U-shaped trajectory: negative values in children (flattening of slope during HV), which increases until adolescence onset (~12 years) becoming positive (steepening during HV), then decreases back, returning negative in adulthood and stabilizing around age ~30 years. The pattern is consistent across brain lobes. **(C)** Comparison of aperiodic slope reactivity for control, seizure, and schizophrenia groups. Trajectories are compared for 5th-95th percentile age range of clinical groups and are shown on linear age scale. Seizure disorder reactivity values are higher than control; schizophrenia trajectories are lower. Since control values are negative in the examined age range, this implied that the seizure group shows hyporeactivity (blunted flattening) while schizophrenia shows hyperreactivity (excessive flattening). **(D)** Effect size (Cohen’s d) of distribution of deviations in individual aperiodic slope reactivities in clinical groups. Orange distributions show positive weak effects for seizure group indicating hyporeactivity (blunted flattening of slope, suggesting compensation); violet distributions show negative weak effects for schizophrenia indicating hyperreactivity.

The clinical reactivity pattern was directionally inverted relative to oscillatory power. Seizure disorder showed hyporeactivity with blunted slope flattening during HV, while schizophrenia showed hyperreactivity with excessive slope flattening, with modest effect sizes (Cohen’s *d <* 0.2) in opposing directions across brain lobes (Fig. 6**C**,**D**). This directional inversion relative to oscillatory power is notable: seizure disorder, which showed oscillatory hyperreactivity, showed blunted broadband E/I shifts, while schizophrenia showed the reverse. Notably, occipital lobe effect sizes were near zero in both groups, indicating regionally preserved aperiodic slope modulation regardless of diagnosis (Fig. 6**D**).

### Inverted-U-Shaped Developmental Aperiodic Offset Reactivity Trajectory Shows Blunted Reductions in Seizure and Schizophrenia

Finally, we examined aperiodic offset reactivity. While aperiodic slope captures the frequency distribution of power as an index of E/I balance, offset reflects total broadband power and indexes overall neural population activity magnitude. Aperiodic offset was consistently lower during HV than rest throughout the lifespan, indicating a broad suppression of neural population activity under metabolic challenge. Offset reactivity followed an inverted U-shaped trajectory: highly negative in childhood, peaking in adolescence while remaining negative, then declining to stable values by young adulthood (~30 years), consistent across all brain lobes (Fig. 7**A**,**B**). The adolescent peak, where the suppressive response to HV is greatest, aligns with inflection points observed across oscillatory and aperiodic slope parameters.

**Figure 7.**
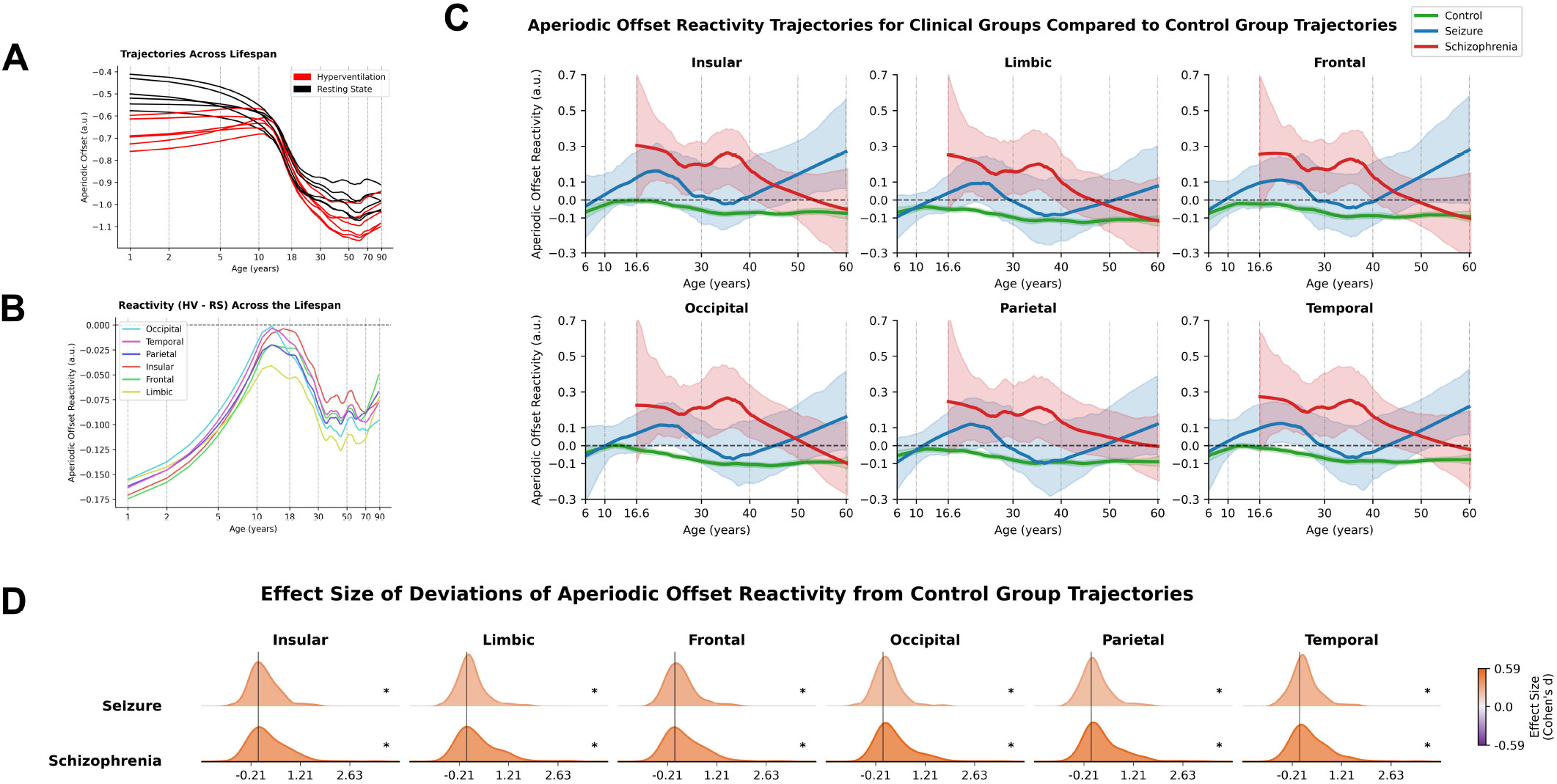
Effects of age and brain pathology on aperiodic offset reactivity: age-related inverted-U-shaped aperiodic offset reactivity shows blunted response in both seizure disorder and schizophrenia. Panels A-D characterize aperiodic offset reactivity across lifespan and clinical groups, stratified by brain lobes. **(A)**Visual comparison of aperiodic offset changes throughout the lifespan for both HV and RS conditions. The age on the x-axis is represented using natural logarithmic scale. Aperiodic offset remains lower during HV compared to RS throughout the lifespan. **(B)** Lifespan reference charts for aperiodic offset reactivity to HV (HV-RS). The age on the x-axis is represented using natural logarithmic scale. Aperiodic offset reactivity shows an inverted-U shaped developmental pattern: starting with large negative values in children, followed by an increase until adolescence while still remaining negative, and then started decreasing until young adulthood (age ~ 30 years), where it stabilizes for the remaining lifespan. The effect is consistent across brain lobes. **(C)** Visual comparison of aperiodic offset reactivity trajectories and the corresponding 95 percent confidence intervals for control, seizure, and schizophrenia groups. The trajectories are shown on a linear age distribution on x-axis. Offset reactivity values for both seizure and schizophrenia groups were higher than the control group. Since the control group trajectories were negative, this implied that both the clinical groups have hyporeactive aperiodic offsets patterns. **(D)** Effect size of distribution of deviations in individual aperiodic offset reactivities in clinical groups. Orange distributions represent positive effects. Statistical significance is marked by (*∗*) (*p <* 0.1). Both seizure and schizophrenia groups demonstrated a shared response in terms of aperiodic offset reactivity with a large positive effect size and statistical significance (*p*¡0.1). The effects are consistent across brain lobes.

The clinical findings were qualitatively distinct from all preceding results. Both seizure disorder and schizophrenia showed hyporeactivity with insufficient suppression of broadband population activity during HV relative to controls, yielding large positive effect sizes (Cohen’s *d >* 0.5) that were statistically significant (*p <* 0.1) across all brain lobes in both groups simultaneously (Figure 7**C**,**D**). This convergence is the central finding of the study. Whereas all oscillatory parameters and aperiodic slope produced modest, divergent effects between the two disorders (Cohen’s *d <* 0.2), aperiodic offset produced large, convergent effects in the same direction despite the opposing pathophysiology of these conditions. This dissociation indicates that alterations in aperiodic offset reactivity are a global and robust phenomenon in brain pathology.

## Discussion

In this study, using large scale routine clinical EEGs, we established lifespan reference charts for EEG spectral parameter reactivity to HV and quantified how neurological and psychiatric disorders alter these responses. We observed that children showed the greatest reactivity compared to adults, with late childhood to adolescence marking a critical transition period for stabilization of EEG spectral power reactivities. Seizure disorder and schizophrenia showed opposite effects on oscillatory EEG reactivities, with seizure disorder showing hyperreactivity in oscillatory power parameters and schizophrenia showing hyporeactivity, consistent with their respective states of neural network hyperexcitability and rigidity. Despite these opposing patterns, both disorders shared a common vulnerability in aperiodic offset reactivity, showing reduced modulation during HV compared to controls with large effect sizes that exceeded all oscillatory effects. These findings represent the first systematic characterization of EEG spectral parameter reactivity to HV across the human lifespan, and the first demonstration that normative lifespan trajectories can disambiguate opposing pathophysiological signatures in neurological and psychiatric disorders while simultaneously exposing convergent deficits in fundamental mechanisms of neural state regulation.

### Adolescence as a Critical Inflection Point for Neural Stress Response Maturation

The convergence of alpha and beta power reactivities around the age of 12 years and the inverted U-shaped trajectories for aperiodic parameters peaking during adolescence reflect maturation of inhibitory systems and their altered vulnerability to HV-induced metabolic stress. HV induces hypocapnia through CO_2_ elimination, causing cerebral vasoconstriction that substantially reduces cerebral blood flow (Brian Jr (1998)). Simultaneously, respiratory alkalosis elevates blood pH, directly affecting neuronal excitability through modulation of neurotransmitter receptors.

At the neurotransmitter level, alkalosis creates asymmetric effects on excitation and inhibition. NMDA receptors are normally suppressed by extracellular protons and alkalosis relieves this tonic inhibition, potentiating excitatory transmission (Traynelis and Cull-Candy (1990); Sinning and Hübner (2013)). Conversely, GABAergic inhibition is impaired through reduced GABA release probability and altered chloride-bicarbonate homeostasis, which shifts the GABAergic reversal potential toward more depolarized values (Tombaugh and Somjen (1996); Kaila et al. (2014)). This creates a net effect of alkalosis preferentially enhancing excitation while weakening inhibition, shifting E/I balance toward excitation.

Children demonstrate enhanced cerebrovascular reactivity to CO_2_ compared to adults, producing more pronounced hemodynamic responses (Settakis et al. (2003)). This heightened vascular sensitivity, in the context of immature E/I balance mechanisms, may contribute to the strong negative reactivity observed in childhood. The GABAergic inhibitory system undergoes protracted maturation through childhood, with developmental shifts in chloride regulation fundamentally altering how neurons respond to GABAergic input (Ben-Ari (2002); Dzhala et al. (2005)). Parvalbumin interneurons that provide fast inhibition and temporal coordination also reach full functionality during adolescence (Caballero et al. (2014)). The coincidence of pH-induced GABAergic disruption with immature inhibitory networks is consistent with the reduced alpha and beta power reactivity observed in childhood.

The adolescent peak in aperiodic reactivity coincides with synaptic pruning that preferentially targets excitatory connections, shifting E/I balance toward inhibition, and concurrent myelination acceleration increasing temporal precision (Petanjek et al. (2011); Paus (2010)). This transition state of maximum neural reorganization combined with maturing but unstabilized inhibitory systems represents maximum dynamic range of neural response but also enhanced vulnerability to neuropsychiatric disorders (Paus et al. (2008)). The distinctive aperiodic signature during adolescence may reflect this unique state of neural flux, consistent with evidence that aperiodic EEG parameters track brain maturation independently of oscillatory measures (Tröndle et al. (2022)).

### Hyperreactive Networks in Seizure Disorders: E/I Imbalance Amplifies Synchronization with Exhausted Homeostatic Capacity

Seizure disorder patients demonstrated hyperreactive alpha and beta power with modest but consistent effect sizes (Cohen’s *d <* 0.2) across brain lobes. The modest effect magnitude likely reflects heterogeneity in epilepsy phenotypes, disease severity, compensatory mechanisms, and anti-seizure medication effects. Frequency modulation remained intact with near-zero effect sizes, revealing dissociation between amplitude and timing control.

The hyperreactive power response is consistent with chronically altered E/I balance favoring excitation, which in epilepsy has been associated with loss of inhibitory interneurons, altered inhibitory receptor function, and enhanced glutamatergic transmission (Fritschy (2008)). Epileptic neurons have lower firing thresholds, causing more neurons to exist in a near-threshold state at baseline (Lévesque et al. (2016)). HV-induced alkalosis, by potentiating NMDA receptors while weakening GABAergic inhibition, may preferentially recruit near-threshold neurons into synchronous activity in epileptic networks. The synchronization machinery, consisting of pyramidal-interneuron feedback loops remains intact or enhanced, potentially amplifying metabolic perturbations into exaggerated oscillatory power responses (Buzsáki and Wang (2012)). For alpha power specifically, this hyperreactivity does not straightforwardly reflect increased inhibitory tone despite alpha oscillations being canonically associated with thalamocortical inhibition. Rather, elevated alpha power in epileptic networks can indicate reduced alpha-related inhibition of cortical regions with abnormal thalamocortical connectivity (Vaudano et al. (2017)), and spike-wave discharges in generalized epilepsies entail hyper-synchronous inhibition in thalamocortical circuits (Shao et al. (2019)), demonstrating that pathological alpha-range synchrony can paradoxically coexist with network hyperexcitability. Enhanced alpha reactivity to HV in epilepsy thus likely reflects aberrant thalamocortical entrainment under metabolic stress rather than a healthy upregulation of inhibitory tone.

Preserved frequency modulation despite amplitude hyperreactivity reveals that frequency is determined by intrinsic membrane properties and synaptic kinetics relatively independent of recruitment processes (Pauluis et al. (1999); Brunel and Wang (2003)). However, specific mechanisms underlying this dissociation remain incompletely characterized in the hyperventilation literature, requiring validation (Guaranha et al. (2005)).

The paradoxically hyporeactive aperiodic slope suggests an active homeostatic compensation. Aperiodic slope is a proxy of E/I balance, with flatter slopes indicating increased excitation (Gao et al. (2017)). Chronic epilepsy triggers homeostatic adaptations including upregulated inhibitory receptor expression and inhibitory axon sprouting (Brooks-Kayal et al. (1998)). During HV, these compensatory mechanisms may partially counteract expected E/I shifts, which could be reflected in the reduced spectral flattening observed in seizure disorder. Additionally, altered chloride regulation in epileptic tissue shifts the GABAergic reversal potential toward less hyperpolarized values, reducing capacity for further modulation when alkalosis perturbs the chloride-bicarbonate equilibrium (Pallud et al. (2014)), reflecting exhausted dynamic range for E/I modulation.

### Neural Inflexibility in Schizophrenia: NMDA Receptor Hypofunction Disrupts Coordinated Network Responses

Schizophrenia patients demonstrated hyporeactivity across oscillatory measures with modest but consistent effect sizes (Cohen’s *d <* 0.2). The modest magnitude likely reflects clinical heterogeneity ranging from schizotypal disorder to severe schizophrenia and the effects of antipsychotic medication. The consistent hyporeactivity reveals fundamental deficits in adaptive network responses, reflecting neural inflexibility rather than hyperexcitability.

The oscillatory hyporeactivity is consistent with disrupted synchronization associated with NMDA receptor hypofunction (Coyle (2012)). Parvalbumin interneurons express high NMDA receptor levels and depend on NMDA-mediated excitation for function. In schizophrenia, these interneurons show reduced GABA synthesis, reduced parvalbumin levels, and altered electrophysiology that impairs high-frequency firing (Hashimoto et al. (2003); Lewis et al. (2012)). This is associated with impaired capacity to provide synchronized inhibitory pulses coordinating pyramidal cell firing into coherent oscillations (Rotaru et al. (2011)).

Critically, NMDA receptors are normally potentiated by alkalosis. During HV, pH-induced potentiation should enhance NMDA currents, contributing to oscillatory responses. However, in schizophrenia, this potentiation operates on already hypofunctional NMDA receptors. Neural circuits with reduced NMDA reserve may be less able to respond appropriately, consistent with the blunted power and frequency responses observed here. NMDA receptor activation normally modulates oscillation frequency through effects on spike timing (Carlen et al. (2012)). The contrast between impaired frequency modulation in schizophrenia and preserved modulation in seizure disorder is consistent with NMDA function playing a role in flexible frequency control.

The exaggerated aperiodic slope response alongside blunted oscillatory response reflects a disconnect between synchronized and asynchronous neural activity. Parvalbumin interneuron dysfunction, by reducing inhibitory control over pyramidal neurons, may contribute to hyperexcitability during HV-induced alkalosis without sufficient coordination into coherent rhythmic oscillations, consistent with the observed pattern of excessive aperiodic slope flattening coexisting with suppressed oscillatory power (Homayoun and Moghaddam (2007)).

Unlike seizure disorder showing compensatory GABA upregulation, schizophrenia demonstrates failed homeostatic plasticity (Crabtree and Gogos (2014)). NMDA hypofunction during critical developmental periods has been associated with disrupted parvalbumin interneuron maturation and impaired formation of compensatory circuits (Carlen et al. (2012)). Oxidative stress may further limit compensation through its association with damage to metabolically demanding parvalbumin interneurons (Steullet et al. (2017)). Together, these factors are consistent with circuits that may not have developed normal compensatory capacity.

### Aperiodic Offset Hyporeactivity as Shared Vulnerability: Convergent Metabolic Dysfunction

The most striking finding was convergent hyporeactive aperiodic offset in both disorders, with strong effect sizes (Cohen’s *d >* 0.5) and statistical significance (*p <* 0.1) across all brain lobes. This shared vulnerability occurred despite opposite slope patterns reflective of opposite E/I imbalances, pointing toward fundamental deficits in mechanisms regulating baseline neural population activity.

Aperiodic offset reflects total broadband power, correlating directly with population spiking rates (Manning et al. (2009); Miller et al. (2014)). While aperiodic slope captures activity distribution across frequencies (E/I balance), aperiodic offset captures overall activity magnitude. Evidence that respiration phase-locks with aperiodic signal fluctuations and directly influences cortical E/I balance (Kluger et al. (2023)) suggests that HV, as a respiratory maneuver, should produce measurable aperiodic modulation. Indeed, in the control group, HV reduces the aperiodic offset through convergent mechanisms. Hypocapnia-induced vasoconstriction decreases oxygen and glucose delivery to the brain. Respiratory alkalosis hyperpolarizes thalamocortical neurons, reducing their baseline firing rate and cortical drive (Williams et al. (2007)). Brainstem arousal systems show decreased activity, lowering global arousal levels. Finally, reduced metabolic substrate availability may limit energy-intensive synaptic transmission (Harris et al. (2012)).

The hyporeactive offset in both disorders indicates impaired capacity to modulate baseline population activity during metabolic stress. This shared phenotype may reflect different proximal mechanisms converging on astrocyte-neuron metabolic coupling and mitochondrial vulnerabilities.

Astrocytes perform essential metabolic stress response functions: regulating extracellular potassium through spatial buffering (Kofuji and Newman (2004)), clearing glutamate to prevent excitotoxicity (Anderson and Swanson (2000)), buffering pH to moderate alkalosis responses (Chesler (2003)), and providing lactate as energy substrate through the astrocyte-neuron lactate shuttle (Pellerin and Magistretti (2012)). In epilepsy, astrocytes show impaired potassium buffering, reduced glutamate transporter expression, reduced glutamine synthesis enzyme activity, and reactive gliosis representing pathological rather than functional responses (Heuser et al. (2012); Proper et al. (2002); Eid et al. (2004); Steinhäuser et al. (2016); Seifert et al. (2006)). In schizophrenia, astrocyte pathology shows decreased astrocyte numbers in prefrontal cortex, reduced glutamate transporter expression, and impaired metabolic coupling with reduced lactate provision (Rajkowska et al. (2002); Bauer et al. (2008); Martins-de Souza et al. (2009); de Oliveira Figueiredo et al. (2022)). While the patterns differ across the two clinical groups, reactive gliosis versus reduced density, both are associated with compromised astrocyte-neuron metabolic coupling that may limit appropriate metabolic adjustment during HV. Beyond pre-existing pathology, HV-induced cerebral hypoperfusion may further compromise astrocyte function acutely, as reduced oxygen and glucose delivery exacerbates impaired potassium buffering and glutamate clearance in already vulnerable tissue (Pellerin and Magistretti (2012); Brian Jr (1998)), potentially amplifying the offset hyporeactivity observed in clinical groups relative to controls.

Mitochondrial dysfunction provides another convergent mechanism. Both schizophrenia and epilepsy show evidence of impaired mitochondrial function (Manji et al. (2012)). In schizophrenia, widespread mitochondrial function alterations include reduced electron transport chain complex activity and elevated oxidative stress (Prabakaran et al. (2004); Rajasekaran et al. (2015)). In epilepsy, the relationship is bidirectional: primary mitochondrial disorders frequently cause seizures, while seizures cause mitochondrial damage through calcium overload and oxidative stress (Khurana et al. (2008); Zsurka and Kunz (2015)), both showing reduced ATP production capacity. Pre-existing mitochondrial deficits may reduce the energy reserve available to modulate baseline activity during HV stress, resulting in a “metabolically locked” system.

Together, the astrocyte and mitochondrial evidence suggests that both disorders show offset hyporeactivity consistent with different proximal constraints converging on shared metabolic vulnerabilities. In epilepsy, compensatory inhibitory upregulation may establish a functional baseline that limits the range of downward modulation available during metabolic challenge, while metabolic deficits further limit the capacity to reduce baseline activity. In schizophrenia, the constraint is primarily metabolic where NMDA hypofunction and parvalbumin interneuron deficits are associated with disinhibited network states, while impaired astrocyte-neuron coupling and mitochondrial dysfunction may limit appropriate activity reduction during metabolic stress (de Oliveira Figueiredo et al. (2022)). This dual-constraint framework is consistent with the convergent phenotype: both disorders show impaired offset reduction, potentially through different combinations of homeostatic constraint and metabolic limitation.

The large effect sizes for offset compared with smaller effect sizes for oscillatory parameters suggest that aperiodic offset taps more fundamental, less compensated vulnerabilities. This pattern aligns with the concept of equifinality in developmental psychopathology, where diverse pathways converge on common phenotypes (Cicchetti and Rogosch (1996)). The relationship between aperiodic offset and low-frequency neural activity provides additional insight. As the aperiodic 1/f component contributes disproportionately to spectral power at lower frequencies, changes in offset are most prominently expressed in the delta and theta frequency ranges, not as modulation of true oscillatory delta/theta activity, but as shifts in the non-oscillatory broadband baseline upon which these rhythms sit. Delta and theta oscillations are themselves generated through large-scale state transitions depending on neurovascular coupling and metabolic regulation (Steriade et al. (1993)), processes that show evidence of dysfunction in both disorders. Altered low-frequency power during HV in both disorders (Yamatani et al. (1995); Bose et al. (2016)) may therefore reflect a combination of impaired aperiodic baseline modulation and genuinely disrupted slow oscillatory dynamics, both consistent with impaired coordination of large-scale brain functional states

The convergent metabolic vulnerability suggested by offset hyporeactivity raises the question of whether metabolism-targeted interventions might benefit patients across diagnostic categories. Ketogenic diets show established efficacy in drug-resistant epilepsy, with children up to three times more likely to achieve seizure freedom compared to controls (Martin-McGill et al. (2018); Rho (2017)). Emerging evidence suggests similar benefits in schizophrenia and other psychiatric conditions, with pilot trials showing significant psychiatric symptom improvement alongside metabolic benefits (Sethi et al. (2024); Palmer et al. (2019); Palmer (2025)). If convergent metabolic vulnerabilities underlie offset hyporeactivity, interventions targeting astrocyte function, mitochondrial biogenesis, or metabolic flexibility may normalize stress responses across diagnostic categories. If aperiodic offset reactivity proves to be a reliable index of metabolic reserve, it could serve a dual translational function: identifying patients most likely to benefit from metabolism-targeted interventions regardless of primary diagnosis, and providing a measurable endpoint for tracking therapeutic response. This would represent a shift from disorder-specific to mechanism-specific treatment stratification in clinical neurophysiology.

### Limitations and Future Directions

Our control group comprised outpatient recordings rather than strictly healthy participants. While our large sample size enabled averaging individual pathological effects for population trajectories, clinical heterogeneity may have influenced our EEG spectral estimates. Further, weak oscillatory effect sizes require cautious interpretation. Medication effects represent equally important confounds: antiepileptic drugs alter GABAergic transmission, and antipsychotics affect dopaminergic/serotonergic transmission, potentially modifying hyperventilation responses.

A methodological consideration specific to the schizophrenia findings concerns differential hyperventilation effort between groups. Since HV-induced EEG changes are closely tied to breathing effort (Guaranha et al. (2005)), reduced compliance or effort during the maneuver in schizophrenia patients could contribute to the observed hyporeactivity independent of underlying neural mechanisms. Our retrospective clinical dataset did not include measures that would allow effort quantification, limiting our ability to fully disentangle effort-related from pathophysiology-driven effects. Nevertheless, the hyporeactive pattern observed here is consistent with the broader characterization of schizophrenia as exhibiting general physiological unresponsiveness to HV (RUBIN (1942)), and aligns mechanistically with established NMDA receptor hypofunction and parvalbumin interneuron dysfunction. Future studies incorporating prospective effort monitoring would strengthen causal interpretation of blunted spectral responses in this population.

Further, source reconstruction relied on a standardized fsaverage template with uniform application across all age groups, as individual MRIs were unavailable for this large retrospective cohort. This approach may introduce greater spatial uncertainty in younger participants whose head geometry deviates more substantially from the adult template, potentially affecting the precision of lobe-level source estimates in pediatric age ranges.

The mechanistic interpretations of response of clinical groups based on the normative trajectories require direct validation. Simultaneous measurement of cerebral blood flow, tissue pH, and EEG during HV could test whether vascular and pH changes mediate observed patterns. Animal models with specific circuit manipulations could establish causal relationships. Future studies could examine astrocyte function using MR spectroscopy, assess mitochondrial function using phosphorus MRS, and test whether metabolic interventions normalize HV responses, validating offset reactivity as a biomarker for metabolic vulnerability.

Prospective validation studies should test whether offset hyporeactivity predicts treatment response to metabolism-targeted interventions such as ketogenic diet. Extension to other disorders with suspected metabolic dysfunction (bipolar disorder, major depression, autism spectrum disorders) would test the generalizability of offset as a trans-diagnostic biomarker. Additionally, longitudinal studies tracking offset reactivity across the course of brain pathology could determine whether this measure reflects trait vulnerability or state-dependent dysfunction.

## Conclusion

This study established the first lifespan reference charts for EEG spectral parameter reactivity to hyperventilation, providing a normative framework that enables differentiation of pathological from age-typical responses across development. Applied to two disorders with opposing circuit-level pathophysiology, these charts revealed a dissociation in the architecture of dysfunction: oscillatory parameters captured the expected divergence between seizure disorder and schizophrenia with weak effect sizes, while aperiodic offset exposed an unexpected convergence with large effect sizes across all brain lobes. The convergence of two disorders with opposite circuit-level pathophysiology on a shared aperiodic deficit suggests that offset captures a more fundamental level of neural vulnerability than circuit-specific oscillatory signatures. If validated prospectively, offset hyporeactivity could serve as a transdiagnostic biomarker of metabolic reserve capacity, enabling mechanism-specific rather than disorder-specific treatment stratification. Future work should test whether offset reactivity predicts response to metabolism-targeted interventions and extend this framework to additional neuropsychiatric conditions with suspected metabolic dysfunction.

## Data Availability

All data produced in the present study are available upon reasonable request to the authors. Only lobe averaged data will be made available.

## Data availability

Data supporting the findings of this study include de-identified participant-level, lobe-wise averaged values for several spectral parameters analyzed in the manuscript. During peer review, these data and the associated code for the analyses will be made available to editors and reviewers upon request. After publication, access to the minimum dataset will be provided upon reasonable request to the corresponding author, subject to applicable ethics approvals and data-sharing agreements required for human participant data.

## Abbreviations

EEG: 
HV: 
E/I: 
ROI: 
LOWESS: 

